# The major predictors of testing positive for COVID-19 among symptomatic hospitalized patients

**DOI:** 10.1101/2020.09.11.20192963

**Authors:** Samson Barasa, Josephine Kiage-Mokaya, Geraldine Luna, Michael Friedlander

## Abstract

**Introduction:** Increasing corona virus disease 2019 (COVID-19) pre-test probability can minimize testing patients who are less likely to have COVID-19 and therefore reducing personal protective equipment and COVID-19 testing kit use. The aim of this study was to identify patients who were likely to test positive for COVID-19 among symptomatic patients suspected of having COVID-19 during hospitalization by comparing COVID-19 positive and negative patients.

**Method:** We conducted a retrospective chart review of patients who were ≥18 years old and underwent COVID-19 Polymerase chain reaction test because they presented with symptoms thought to be due to COVID-19. The Poisson regression analysis was conducted after clinical presentation, demographic, medical co-morbidities, laboratory and chest image data was retrieved from the medical records.

**Results:** Charts of 277 and 35 COVID-19 negative and positive patients respectively were analyzed. Dyspnea (61%) was the most common symptom among COVID-19 negative patients, while 83% and 77% COVID-19 positive patients had cough and fever respectively.

COVID-19 positive patients were more likely to present initially with cough [1.082 (1.022 - 1.145)] and fever [1.066 (1.013 - 1.122)], besides being males [1.066 (1.013 - 1.123)] and 50 to 69 years old [1.090 (1.019 - 1.166)]. Dyspnea, weakness, lymphopenia and bilateral chest image abnormality were not associated with COVID-19 positivity.

COVID-19 positive patients were less likely to have non-COVID-19 respiratory viral illness [0.934 (0.893 - 0.976)], human immunodeficiency virus [0.847 (0.763 - 0.942)] and heart failure history [0.945 (0.908 - 0.984)]. Other chronic medical problems (hypertension, diabetes mellitus, chronic obstructive pulmonary disease and coronary artery disease) were not associated with testing positive for COVID-19.

**Conclusion:** Cough and fever are better predictors of symptomatic COVID-19 positivity during hospitalization. Despite published studies reporting a high prevalence of lymphopenia among COVID-19 positive patients, lymphopenia is not associated with the risk of testing positive for COVID-19.

## Introduction

The center for disease control (CDC) has recommended corona virus 2019 (COVID-19) testing for patients who have fever and or respiratory symptoms such as cough and dyspnea among hospitalized patients, healthcare workers and elderly patients, besides those exposed to COVID-19 and those in high prevalence COVID-19 communities^1^.

Patients with chronic obstructive pulmonary disease (COPD),^2^ asthma^3^ or congestive heart failure (CHF)^4^ exacerbation can present with cough and dyspnea; while patients with bacterial pneumonia, non-COVID-19 viral pneumonia, bronchitis or upper respiratory tract infections^5^ can present with fever besides cough and dyspnea.

The initial COVID-19 publication from Mainland, China found that 83 % COVID-19 positive patients had lymphopenia and 58.3% had bilateral chest image abnormality.^6^ Subsequent studies have also reported a high lymphopenia prevalence among COVID-19 positive patients.^7,8^ However, these published studies did not compare the prevalence of lymphopenia and or bilateral chest image abnormality among COVID-19 positive and negative patients presenting with cough, dyspnea and or fever in the hospital.

Our study sought to determine whether lymphopenia and or bilateral chest image abnormality in association with cough, dyspnea, fever or weakness increased the risk of testing positive for COVID-19. Increasing the COVID-19 pre-test probability will minimize testing patients who are less likely to have COVID-19 and therefore reducing personal protective equipment (PPE) and COVID-19 testing kit use. In addition, increasing the pre-test probability will further increase the positive predictive value of COVID-19 polymerase chain reaction (PCR) tests.

## Methodology

### Study population and setting

A retrospective chart review of each consecutive patient who was tested for COVID-19 using the PCR test after presenting in the hospital with symptoms thought to be due to COVID-19, between 2^nd^ February 2020 and 29^th^ August 2020 was conducted in Eugene, Oregon. The study enrollees were 18 years and older. The PeaceHealth institutional review board (IRB) approved the study and waived the written informed consent requirement.

### Data collection

The Microsoft Access software was used to manage data after being extracted from the electronic medical records.

### Risk factors

Data was obtained from documents, laboratory tests and chest images completed when the patient initially arrived in the hospital. Information on whether the patient had cough, fever, dyspnea, fatigue or weakness was retrieved from the hospital triage note. Lymphopenia data was gathered from the complete blood count results, while bilateral chest image abnormality was determined by reviewing the radiologist’s chest image report. The non-COVID-19 respiratory viral illness data was collected from the respiratory viral panel results.

The following risk factors were also examined: age, diabetes mellitus, stroke, dementia, CHF, COPD, asthma, systemic lupus erythromatosus (SLE), organ transplant, coronary artery disease (CAD), smoking, hypertension and body mass index (BMI.

### Outcome

The study outcome was the COVID-19 PCR test result.

### Statistical analysis

Our study hypothesized that COVID-19 positive patients were more likely to have lymphopenia and or bilateral chest image abnormality in association with cough, fever, dyspnea or weakness compared to COVID-19 negative patients.

The Generalized Poisson regression analysis was used to determine whether COVID-19 positive patients were more likely to have bilateral chest image abnormality, lymphopenia, cough, fever, dyspnea or weakness adjusting for non-COVID-19 respiratory viral illness, history of smoking, asthma, diabetes mellitus, chronic kidney disease, SLE, COPD, CHF, hypertension and BMI. The statistical analysis was performed using STATA version 15.

## Results

**Table I:**
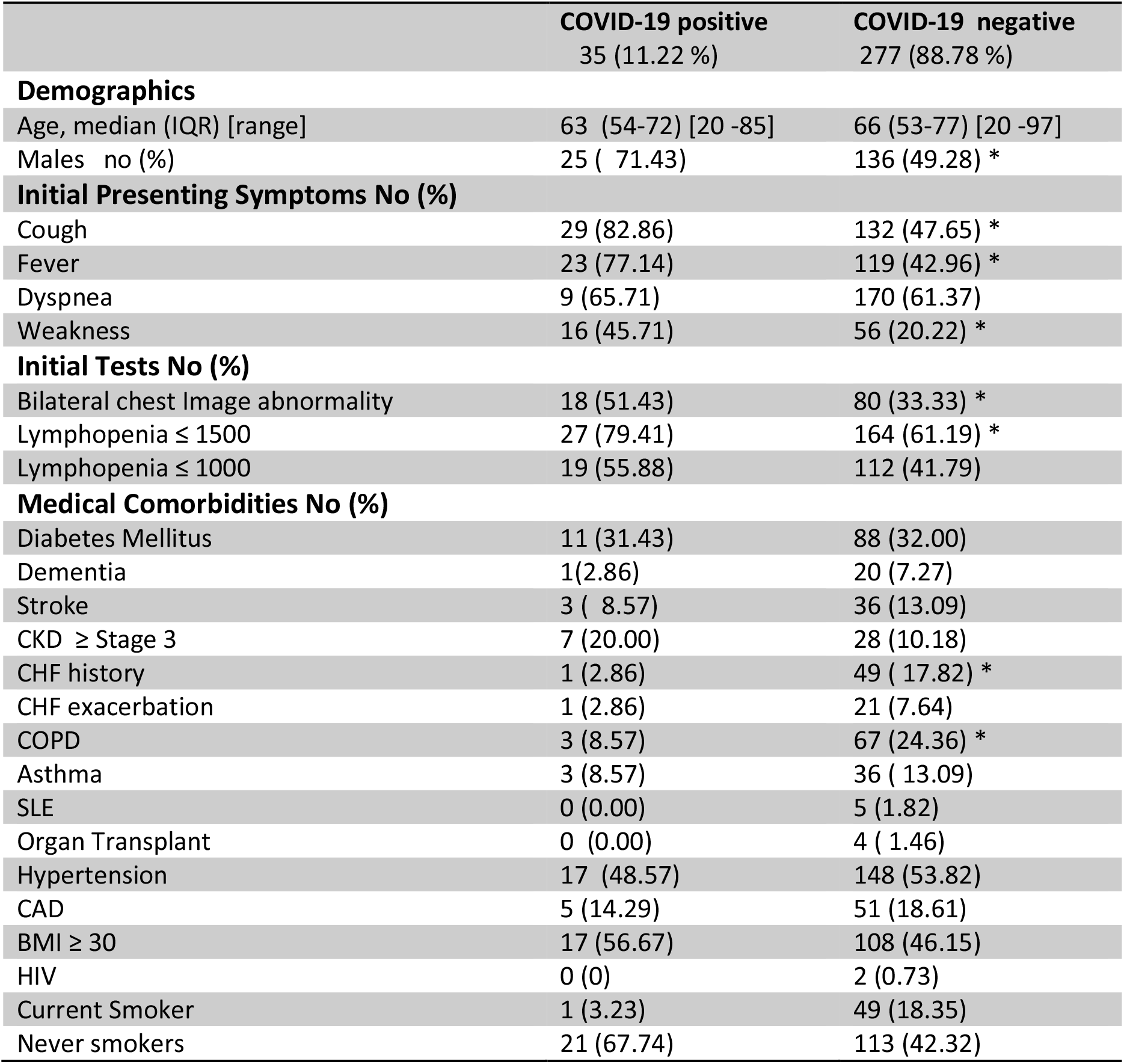
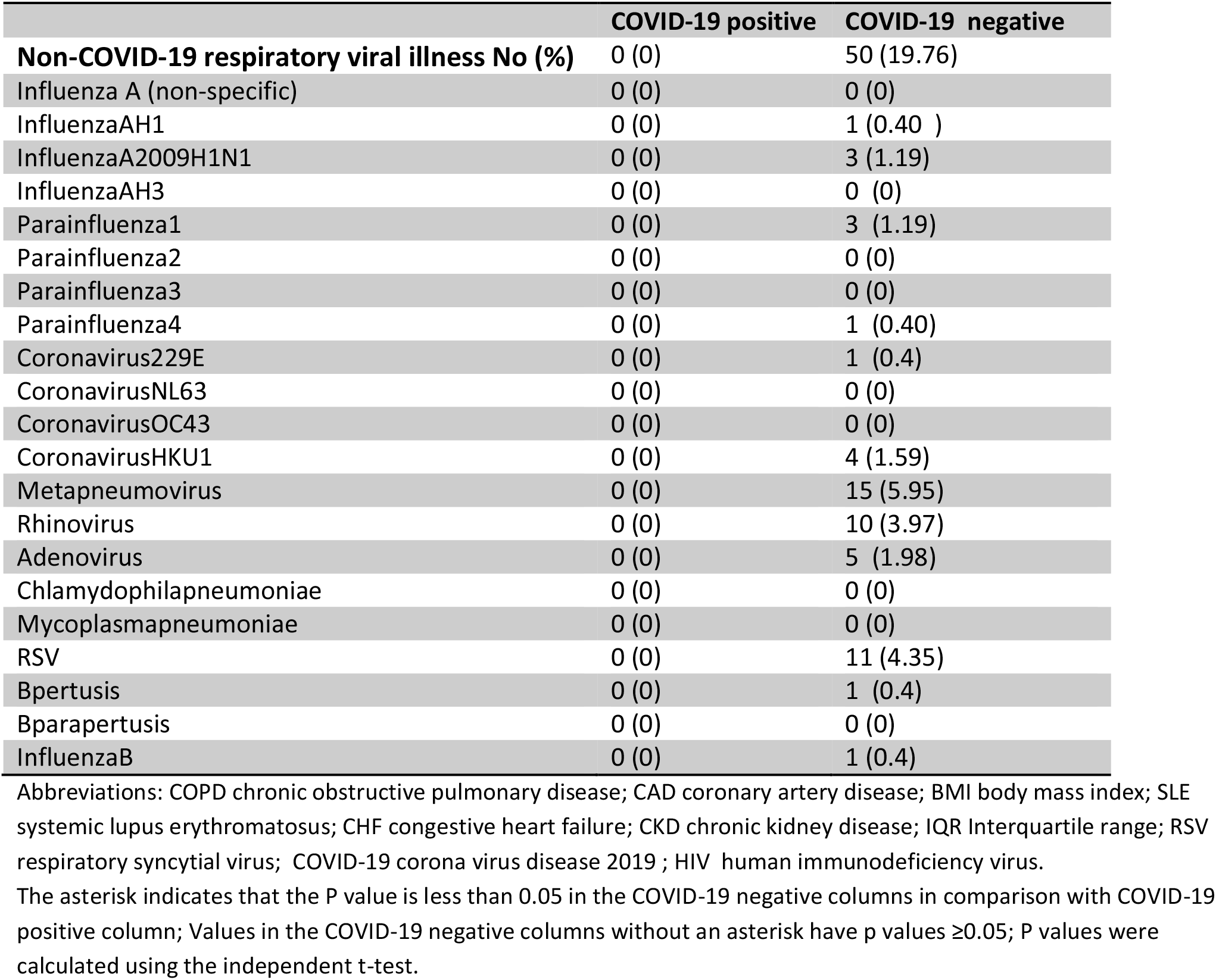
Baseline characteristics of study enrollees.

### Demographics

The median age and the interquartile age range for COVID-19 positive and negative patients was not significantly different, although the age range of COVID-19 positive patients was 20 – 85 years, while the age range of COVID-19 negative patients was 20 to 97 years. Most COVID-19 positive enrollees were males (71%) compared to the equal gender distribution among the COVID-19 negative enrollees.

### Medical comorbidities

The prevalence of medical comorbidities among COVID-19 positive and negative patients was not significantly different apart from CHF and COPD. COVID-19 positive patients had a significantly lower prevalence of CHF and COPD compared to the COVID-19 negative patients. Hypertension was the most common medical comorbidity among COVID-19 positive (49%) and negative (54%) patients. Similarly, diabetes mellitus was the second most common chronic medical problem among COVID-19 positive and negative enrollees.

3% of COVID-19 positive patients were current smokers, while 18% COVID-19 negative patients were current smokers. 67.74 % and 42.3 % COVID-19 positive and negative patients respectively were never smokers.

### Initial Clinical Presentation

Figure 1 below shows the prevalence of the initial clinical features for COVID-19 positive and negative patients during hospitalization. COVID-19 positive patients had a significantly higher prevalence of cough compared to the COVID-19 negative patients. Cough (83%) was the most common symptom followed by fever (77%) and dyspnea (66%) among COVID-19 positive patients. Dyspnea (61.4%) was the most common symptom among COVID-19 negative patients.

**Figure 1:**
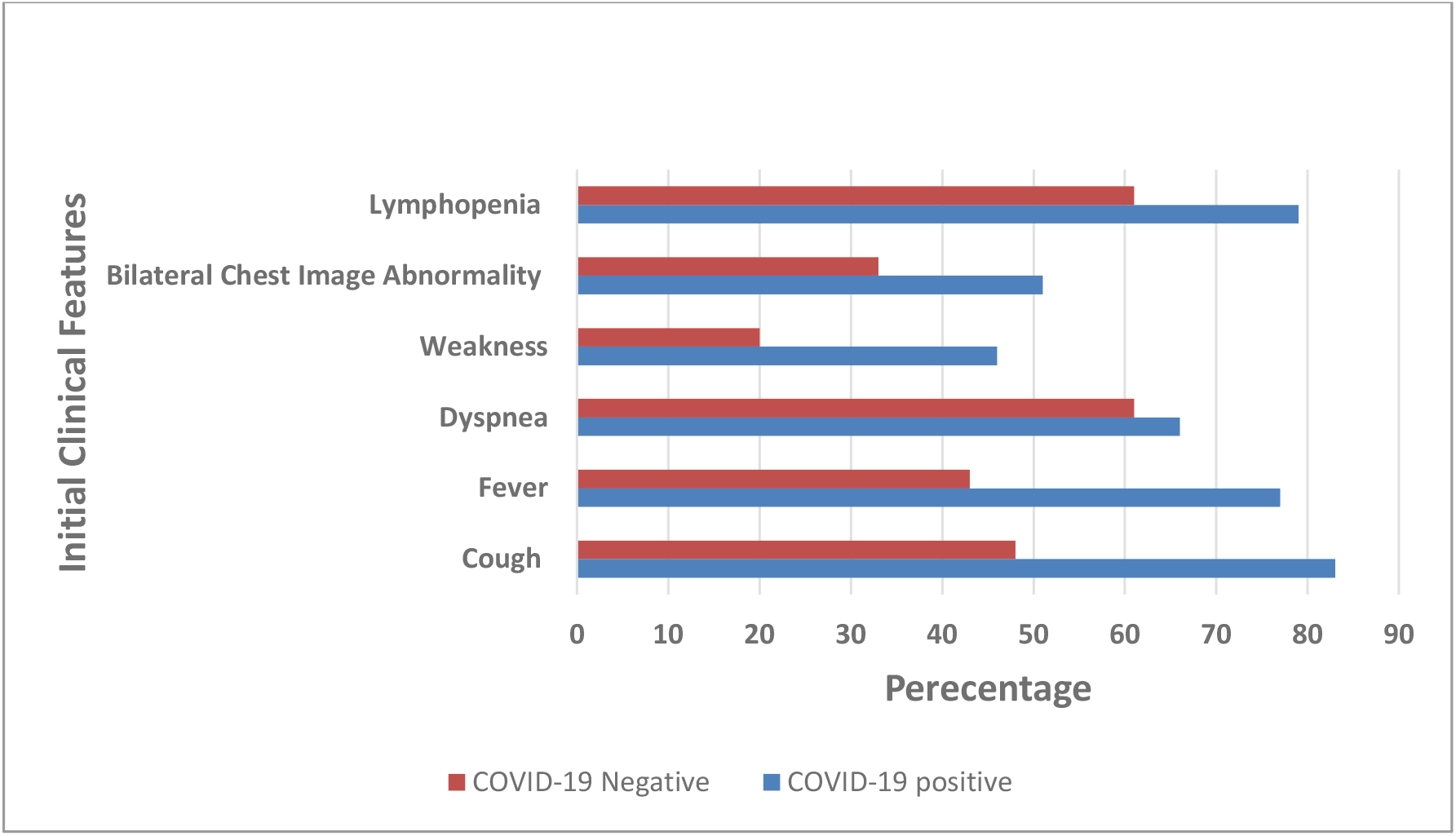
COVID-19 Positive and Negative Patients’ Initial Clinical Features

51% COVID-19 positive patients had bilateral chest image abnormality compared to 33% COVID-19 negative patients. The prevalence of lymphopenia among COVID-19 positive (79%) and negative (61%) patients was not significantly different.

### Associations

**Table 2** below shows the association between COVID-19 status and coughing. COVID-19 positive patients were more likely to present with cough, be 50 to 69 years old and not have non-COVID-19 respiratory viral illness. Moreover, lymphopenia was not associated with COVID-19 positivity. On the other hand, COVID-19 positive patients were less likely to have history of CHF or HIV.

**Table 2:** The Association between COVID-19 Status and Cough

| Table 2: The Association between COVID-19 Status and Cough |  |  |  |  |
| --- | --- | --- | --- | --- |
|  | COVID-19 Status |  | P-value | RR (Confidence Interval) |
| Initial clinical presentation | Cough | Absent | Reference | 1 |
|  |  | Present | 0.007 | 1.082 (1.022 - 1.145) |
| | Lymphopenia ($\leq 1500$) | | 0.809 | 1.006 (0.955 - 1.060) |
| Age Categories | 20 to 39 years (reference) |  |  | 1 |
|  | 40 to 49 years |  | 0.484 | 1.019 (0.966 - 1.076) |
|  | 50 to 69 years |  | 0.012 | 1.090 (1.019 - 1.166) |
|  | 70 to 99 years |  | 0.194 | 1.049 (976 - 1.128) |
| **Medical Comorbidities** | Never Smokers |  | 0.419 | 1.024 (.967 - 1.084) |
|  | Non-COVID-19 respiratory viral illness | Absent | Reference | 1 |
|  |  | Present | 0.003 | 0.934 (0.893 - 0.976) |
|  | Hypertension |  | 0.167 | 1.042 (0.983 - 1.105) |
|  | Diabetes |  | 0.606 | 0.986 (0.933 - 1.041) |
|  | COPD |  | 0.140 | 0.963 (0.915 - 1.013) |
|  | Asthma |  | 0.510 | 0.976 (0.909 - 1.049) |
|  | SLE |  | 0.743 | 0.986 (0.905 - 1.074) |
|  | Organ Transplant |  | 0.086 | 0.908 (0.813 - 1.014) |
| | CKD ≥ Stage 3 | | 0.421 | 1.045 (.939 - 1.163) |
|  | CHF history | Absent | Reference | 1 |
|  |  | Present | 0.006 | 0.945 (0.908 - 0.984) |
|  | HIV | Absent | Reference | 1 |
|  |  | Present | 0.002 | 0.847 (0.763 - 0.942) |
| | BMI ≥ 30 | | 0.617 | 1.001 (0.997 - 1.005) |

As indicated in **Table 4** below, COVID-19 positive patients were more likely to have fever. Besides that, COVID-19 positive patients were more likely to be 50 to 69 years old and less likely to have history of CHF, organ transplant or HIV. Lymphopenia was not associated with COVID-19 status.

**Table 4:**
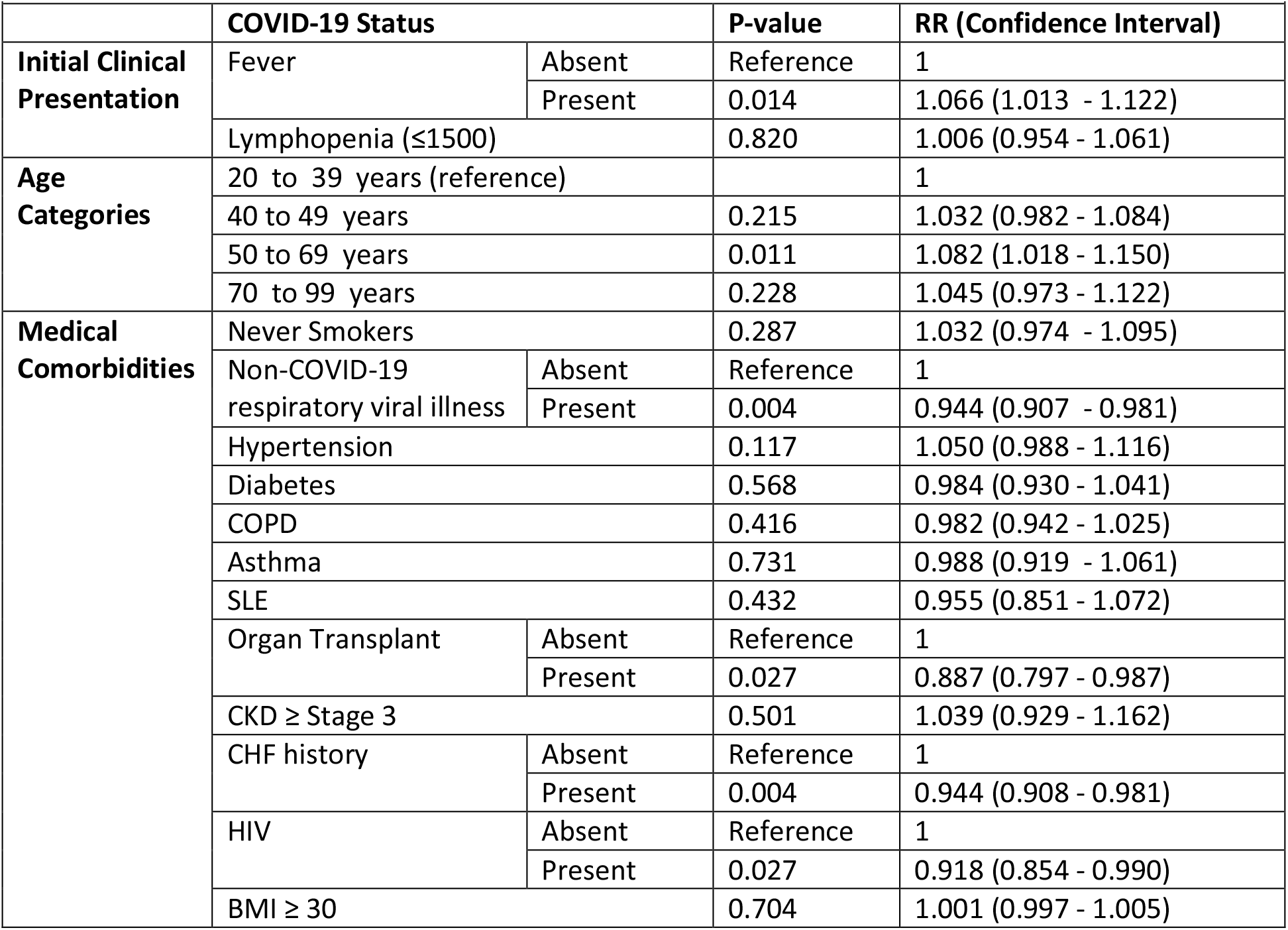
The Association between COVID-19 Status and Fever

The bilateral chest image analysis (**Table 5**) demonstrated that COVID-19 status was not associated with bilateral chest image abnormality and reaffirmed that COVID-19 positive patients were less likely to have non-COVID-19 respiratory viral illness, history of CHF or HIV.

**Table 5:**
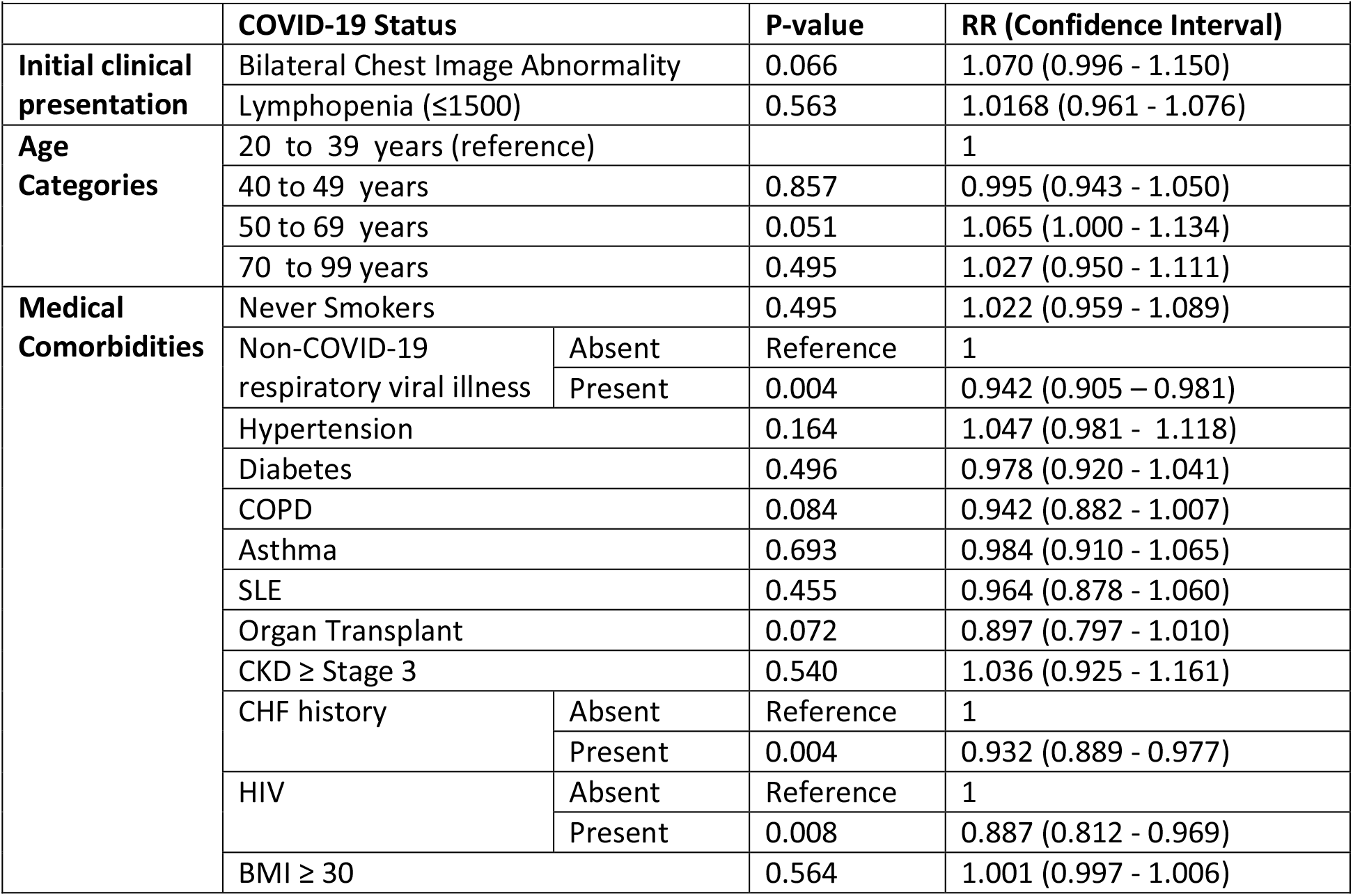
The Association between COVID-19 Status and Bilateral Chest Image Abnormality

Lastly, Males were more likely to test positive for COVID-19. On the other hand, COVID-19 status was not associated with dyspnea [1.013 (0.959 - 1.069)] or weakness [1.054 (0.980 - 1.134)]. For details of these analysis, please see tables 6, 7 and 8 in supplementary I.

## Discussion

### Predictors of COVID-19 positivity

We found that COVID-19 positive patients were more likely to present initially with cough and fever, besides being males and 50 to 69 years old during hospitalization. Furthermore, COVID-19 positivity was not associated with dyspnea, weakness or bilateral chest image abnormality. Despite published studies reporting a high prevalence of lymphopenia among COVID-19 positive patients,^6,7,8^ COVID-19 negative patients too had a high prevalence of lymphopenia. Lymphopenia was not associated with an increased risk of testing positive for COVID-19. Medical comorbidities such as diabetes mellitus, hypertension, coronary artery disease, COPD and SLE didn’t increase the risk of testing positive for COVID-19. However, COVID-19 positive patients were less likely to present with non-COVID-19 respiratory viral illness and have CHF, HIV or history of organ transplant.

### Strengths of this Study

The initial COVID-19 published (Mainland, China;^6^ New York, USA;^9^ and Seattle, USA^8^) studies identified the characteristics of COVID-19 positive patients. However, these studies and subsequent studies^10,11^ didn’t compare patients who tested positive and negative for COVID-19 among patients who presented in the hospital with symptoms thought to be due to COVID-19.

Our study compared the initial clinical presentation of patients who tested positive and negative for COVID-19 among patients who presented in the hospital with symptoms suspected to be due to COVID-19. Although the individualized risk prediction study compared COVID-19 positive and negative patients, it was conducted in Cleveland clinics, included drive-thru sites and didn’t report on chest image abnormality or lymphopenia;^12^ moreover, COVID-19 positive patients in the Cleveland clinics Ohio development cohort (sample 818) had a very low prevalence of cough (15%), fever (13%) and dyspnea (8%) among COVID-19 positive patients.^12^ In contrast, the hospitalized COVID-19 positive patients in our study, the Seattle study, the New York study and the Mainland China study had a high prevalence of cough, fever and dyspnea.^6, 8, 9^

COVID-19 positive patients in our study had comparable demographic and baseline characteristics to the New York study (with 5700 enrollees) at the epicenter of the COVID-19 pandemic in USA.^9^ On the other hand, the mainland China study (with 1099 enrollees) had younger patients with a low prevalence of medical comorbidities.^6^

### Demographics

The interquartile range of COVID-19 positive patients was between 54 and 72 years, while the median age was 63 years in our study. Similarly, COVID-19 positive patients in the New York study had an interquartile range of 52-75 years and a median age of 63 years.^9^ In contrast, patients in the Mainland, China study were younger with a median age of 47 years and an interquartile range of 35 to 58 years.^6^

Our study found that COVID-19 positive patients were more likely to be 50 to 69 years old. In congruence, the Cleveland study found that COVID-19 positivity risk increased with age.^12^ 71% of COVID-19 positive patients in our study and approximately 60% COVID-19 positive patients in the New York and China studies were males.^6,^^9^ Males were at higher risk of testing positive for COVID-19 in our study and the Cleveland study, despite the Cleveland study having an equal gender distribution among COVID-19 positive patients.^12^

### Medical Comorbidities

Our study found that medical comorbidities were not associated with an increased risk of testing positive for COVID-19. The prevalence of medical comorbidities in our study and the New York study was comparable.^9^ The medical comorbidities examined in our study among COVID-19 patients were: hypertension 48.6%, diabetes mellitus 31.4%, CAD 14.3%, COPD 8.6%, CKD 20%, asthma 8.6 %, CHF 3% and BMI (≥ 30) 56.7%. While the prevalence of medical comorbidities analyzed in the New York study was: hypertension 56.6%, diabetes mellitus 33.8%, CAD 11.1%, COPD 5.4%, CKD 5%, asthma 9%, CHF 6.9% and BMI (**≥ 30)** 41.7%.^9^ The Mainland, China study reported a lower prevalence of medical comorbidities, namely: hypertension (15%), diabetes mellitus (7.4%), CAD (2.5%), COPD (1.1%) and CKD (0.7%).^6^ The China study didn’t publish asthma or BMI data.^6^ The China publication and our study found a stroke prevalence of 1.4% and 11.1% respectively among COVID-19 positive patients, while the New York study didn’t report on stroke prevalence.^6, 9^

The prevalence of CHF, HIV and organ transplant among COVID-19 positive patients in the New York study was 0.8% and 1 % respectively;^9^ while none of the COVID-19 positive patients in our study had these medical comorbidities. Moreover, our study found that COVID-19 positive patients were less likely to have CHF, HIV and Organ transplant.

Similarly, COVID-19 positive patients were less likely to have non-COVID-19 respiratory viral illness in our study. The prevalence of non-COVD-19 respiratory viral illness was 0% in our study, while in the New York study it was 2.1%.^9^ The mainland, China study didn’t report the prevalence of non-COVID-19 respiratory viral illness.^6^

### Initial Presenting Symptoms

67%, 43.8%, 18.7% and 38.1% COVID-19 positive patients initially presented with cough, confirmed fever (≥37.5°C), dyspnea and fatigue respectively in the Mainland, China study.^6^ While 30.7% COVID-19 positive patients had confirmed fever (>38°C) in the New York study; however this New York study didn’t report the prevalence of cough, subjective fever, dyspnea, weakness or fatigue.^9^ A study of 393 COVID-19 positive patients conducted in two New York hospitals, with comparable prevalence of medical comorbidities to the aforementioned (larger) New York study and our study, found that the initial COVID-19 positive patients’ symptoms on presentation were cough (79.4%), subjective fever (77.1%) and dyspnea (56.5%), while 25.5% had confirmed fever (> 38°C) on hospital arrival.^10^ Other studies also found that cough was the most common initial COVID-19 presenting symptom.^8, 12,11^

In our study 83 %, 77%, 66% and 46% COVID-19 positive patients had cough, subjective fever, dyspnea and fatigue respectively. Cough was the most common initial symptom among hospitalized COVID-19 patients followed with fever. COVID-19 positive patients were more likely to have cough or fever in our study while dyspnea and weakness were not associated with COVID-19 positivity.

### Chest Image

36.5% of all hospitalized COVID-19 positive patients and 58.3% of those with severe COVID-19 disease had bilateral chest image abnormality during hospital admission in Mainland, China.^6^ On the other hand, the New York study didn’t publish data on chest image abnormality.^9^ However, the New York study with 393 COVID-19 positive enrollees found that 59.8% enrollees had bilateral chest image abnormality during hospital admission.^10^

51.4 % COVID-19 positive patients in our study had bilateral chest image abnormality; however COVID-19 status was not associated with bilateral chest image abnormality.

### Lymphopenia

90%, 73% and 83% COVID-19 positive patients in the New York, Seattle and China studies respectively had a lymphopenia of ≤ 1500 compared to 79.4% in our study.^6, 8, 10^ The New York study and our study found that 60% and 56% COVID-19 positive patients respectively had a lymphopenia of ≤ 1000.^9^ Although the prevalence of lymphopenia was significantly high among COVID-19 positive patients compared to the COVID-19 negative patients in our study, lymphopenia was not associated with COVID-19 positivity.

### Limitations

Limitations of our study included missing data and not all patients had laboratory and chest image tests done. Non-COVID-19 respiratory viral illness data was missing for some patients, because the hospital stopped testing for non-COVID-19 respiratory viral illness in the middle of the pandemic. In addition, we didn’t have data on how long those with history of smoking had quit smoking as well as whether their presenting cough symptom was new, an exacerbation or their baseline smokers’ cough.

## Conclusion

Based on this study, cough and fever are better predictors of testing positive for COVID-19 among patients presenting in the hospital with symptoms thought to be due to COVID-19. Moreover, being 50 to 69 years old and a male increases the risk of testing positive for COVID-19, while bilateral chest image abnormality is not associated with COVID-19 status.

The findings of this study also suggest that COVID-19 positive patients are less likely to have non-COVID-19 respiratory viral illness, HIV or CHF.

Despite published studies reporting a high prevalence of lymphopenia among COVID-19 positive patients, lymphopenia doesn’t increase the risk of COVID-19 positivity. Lastly, non-lymphopenic patients and those without medical comorbidities are not at a lower risk of testing positive for COVID-19.

## Data Availability

all data referred to in the manuscript is available in the manuscript.

## Supplementary 1 Table 6

**Table 6:**
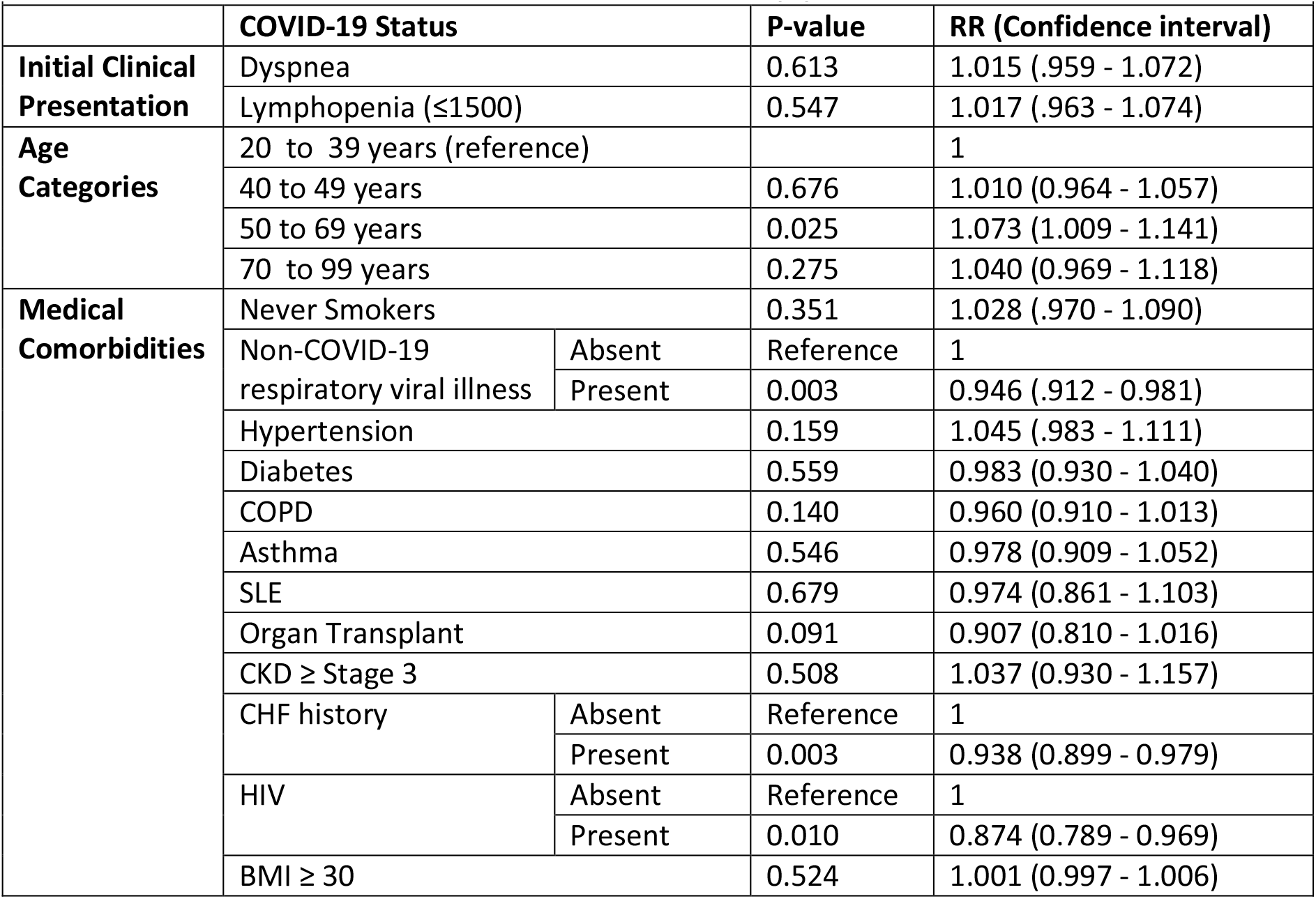
The Association between COVID-19 Status and Dyspnea

### Supplementary 1 Table 7

**Table 7:**
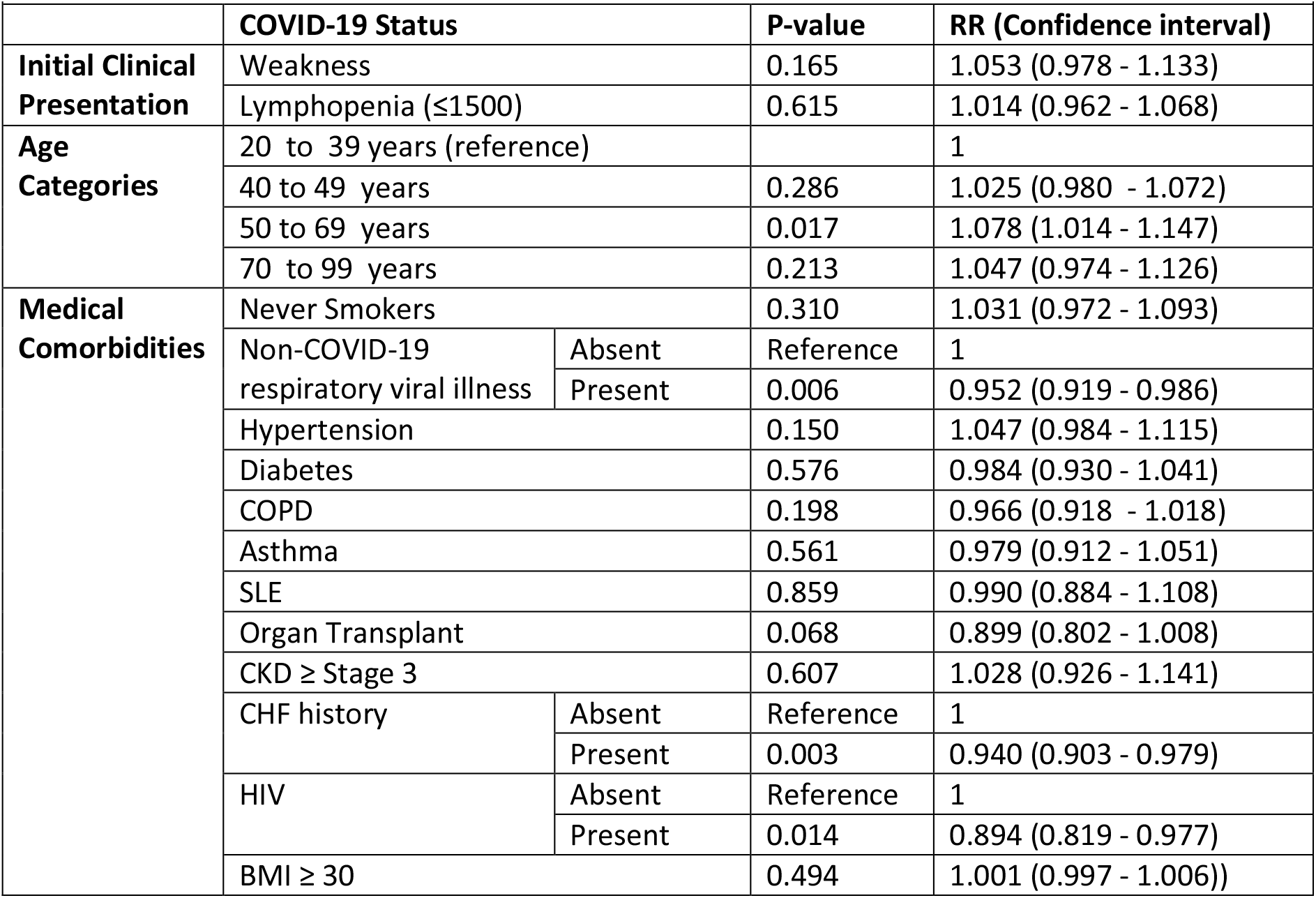
The Association between COVID-19 Status and Weakness

| Table 7: The Association between COVID-19 Status and Weakness |  |  |  |  |
| --- | --- | --- | --- | --- |
|  | COVID-19 Status |  | P-value | RR (Confidence interval) |
| Initial Clinical Presentation | Weakness |  | 0.165 | 1.053 (0.978 - 1.133) |
| | Lymphopenia ($\leq 1500$) | | 0.615 | 1.014 (0.962 - 1.068) |
| Age Categories | 20 to 39 years (reference) |  |  | 1 |
|  | 40 to 49 years |  | 0.286 | 1.025 (0.980 - 1.072) |
|  | 50 to 69 years |  | 0.017 | 1.078 (1.014 - 1.147) |
|  | 70 to 99 years |  | 0.213 | 1.047 (0.974 - 1.126) |
| Medical Comorbidities | Never Smokers |  | 0.310 | 1.031 (0.972 - 1.093) |
|  | Non-COVID-19 respiratory viral illness | Absent | Reference | 1 |
|  |  | Present | 0.006 | 0.952 (0.919 - 0.986) |
|  | Hypertension |  | 0.150 | 1.047 (0.984 - 1.115) |
|  | Diabetes |  | 0.576 | 0.984 (0.930 - 1.041) |
|  | COPD |  | 0.198 | 0.966 (0.918 - 1.018) |
|  | Asthma |  | 0.561 | 0.979 (0.912 - 1.051) |
|  | SLE |  | 0.859 | 0.990 (0.884 - 1.108) |
|  | Organ Transplant |  | 0.068 | 0.899 (0.802 - 1.008) |
| | CKD ≥ Stage 3 | | 0.607 | 1.028 (0.926 - 1.141) |
|  | CHF history | Absent | Reference | 1 |
|  |  | Present | 0.003 | 0.940 (0.903 - 0.979) |
|  | HIV | Absent | Reference | 1 |
|  |  | Present | 0.014 | 0.894 (0.819 - 0.977) |
| | BMI $\geq 30$ | | 0.494 | 1.001 (0.997 - 1.006)) |

### Supplementary 1 Table 8

**Table 8:**
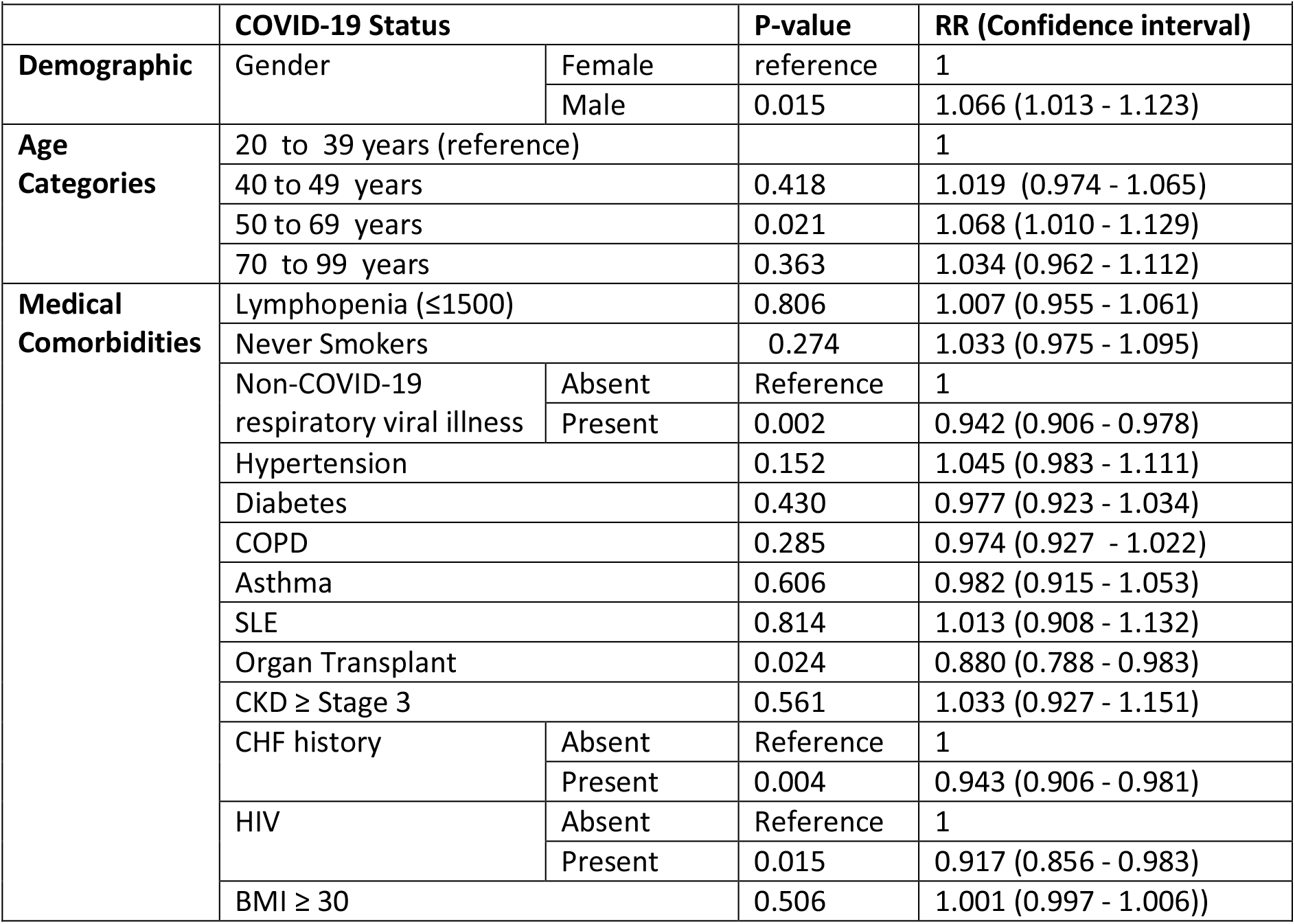
The Association between COVID-19 Status and Gender

## References

1. CDC. Evaluating and Testing Persons for Coronavirus Disease 2019 (COVID-19) Available: https://www.cdc.gov/coronavirus/2019-ncov/hcp/clinical-criteria.html. Accessed 3/21/20, 2020.

2. Qureshi H, Sharafkhaneh A, Hanania NA. Chronic obstructive pulmonary disease exacerbations: latest evidence and clinical implications. Therapeutic advances in chronic disease. 2014; 5(5): 212–227.

3. Quirt J, Hildebrand KJ, Mazza J, et al. Asthma. Allergy, asthma, and clinical immunology: official journal of the Canadian Society of Allergy and Clinical Immunology. 2018;14(Suppl 2): 50.

4. Inamdar AA, Inamdar AC. Heart Failure: Diagnosis, Management and Utilization. Journal of clinical medicine. 2016; 5(7).

5. Banning M. Influenza: incidence, symptoms and treatment. British journal of nursing. 2005; 14(22): 1192–1197.

6. Ding Q, Lu P, Fan Y, et al. The clinical characteristics of pneumonia patients co-infected with 2019 novel coronavirus and influenza virus in Wuhan, China. Journal of medical virology. 2020.

7. Huang I, Pranata R. Lymphopenia in severe coronavirus disease-2019 (COVID-19): systematic review and meta-analysis. Journal of intensive care. 2020; 8: 36.

8. Buckner FS, McCulloch DJ, Atluri V, et al. Clinical Features and Outcomes of 105 Hospitalized patients with COVID-19 in Seattle, Washington. Clinical infectious diseases: an official publication of the Infectious Diseases Society of America. 2020.

9. Richardson S, Hirsch JS, Narasimhan M, et al. Presenting Characteristics, Comorbidities, and Outcomes Among 5700 Patients Hospitalized With COVID-19 in the New York City Area. JAMA: the journal of the American Medical Association. 2020.

10. Goyal P, Choi JJ, Pinheiro LC, et al. Clinical Characteristics of Covid-19 in New York City. The New England journal of medicine. 2020; 382(24): 2372–2374.

11. Jehi L, Ji X, Milinovich A, et al. Development and validation of a model for individualized prediction of hospitalization risk in 4,536 patients with COVID-19. PloS one. 2020; 15(8): e0237419.

12. Jehi L, Ji X, Milinovich A, et al. Individualizing Risk Prediction for Positive Coronavirus Disease 2019 Testing: Results from 11,672 Patients. Chest. 2020.

